# Coronavirus (COVID-19) Spike in Georgia: An Epidemiologic Study of Data, Modelling, and Policy Implications to Understand the Gender-and Race-Specific Variations

**DOI:** 10.1101/2021.12.09.21267571

**Authors:** Fahad Mostafa, Riya Ganji, Julie St. John, Hafiz Khan

## Abstract

**Objective:** The purpose of this study was to investigate the gender-and race-specific predictive variations in COVID-19 cases and deaths in Georgia, USA.

**Methods:** The data were extracted from the Georgia Department of Public Health (GDPH). Statistical methods, such as descriptive statistics, Artificial neural networks (ANN), and Bayesian approach, were utilized to analyze the data.

**Results:** More Whites died from COVID-19 than African-Americans/Blacks in Cobb, Hall, Gwinnett, and non-Georgia residents; however, more Blacks died in Dekalb and Fulton counties. The highest posterior mean for female deaths was obtained in Gwinnett County (77.17; 95% CI, 74.23–80.07) and for male deaths in Fulton County (73.48; 95% CI, 72.18–74.49). For overall race/ethnicity, Whites had the highest posterior mean for deaths (183.18; 95% CI, 128.29–238.27) compared with Blacks (162.48; 95% CI, 127.15– 197.42). Assessing the classification of the chronic medical conditions using ANN, Cobb and Hall Counties showed the highest mean AUC-ROC of the models (78% and 79%, respectively).

**Conclusions:** The predictive models of COVID-19 transmission will help public health practitioners and researchers to better understand the course of the COVID-19 pandemic. The study findings are generalizable to populations with geographic and racial/ethnic similarities and may be used to determine gender/race-specific future virus models for effective interventions or policy modifications.

**Human Subjects:** No personal identifiable information was obtained.

## Introduction

At the end of 2019, a serious airborne viral pandemic, caused by the severe acute respiratory syndrome coronavirus 2 (SARS-CoV-2), rapidly swept across the globe. This coronavirus is part of the *coronaviridae* family of the *Nidovirales* order, a viral family known to cause outbreaks such as the SARS epidemic of 2003 and the MERS epidemic of 2012. Although related to the *coronaviridae* family, SARS-CoV-2 is completely a novel virus of its own and causes the disease known as COVID-19 or coronavirus disease 2019. SARS-CoV-2 infects human hosts by infiltrating alveolar lung cells with a viral spike glycoprotein.^1^ Inside the host, the virus replicates, thrives and then spreads to other hosts through respiratory droplets. SARS-CoV-2 has a high reproductive quotient of 2.24 to 3.58^2^-meaning one person with the disease can infect about 3 additional people on average. The symptoms of COVID-19 vary and include difficulty breathing, chest pain, cough, fever, loss of smell, loss of taste, nausea, vomiting, and more.^3^ The disease has affected the elderly and immunocompromised more severely, and the risk of high severity also increases with comorbid conditions such as diabetes, heart conditions, and obesity.^4^

As of February 27, 2021, COVID-19 has infected over 113 million individuals worldwide and killed over 2.5 million people.^5^ In the United States, there have been over 28 million cases and over 508,000 deaths as of February 27, 2021.^6^ Of these numbers, over 1,000,400 cases and 17,000 deaths have been in the state of Georgia.^6^ This study examined the epidemiology of COVID-19 disease in five Georgia Counties: Cobb, Dekalb, Fulton, Gwinnett, and Hall. Additionally, the study included cases from the non-GA resident group, who are non-Georgian residents diagnosed in Georgia. These groups were selected for study because they have the highest numbers of cases and deaths in Georgia. **Table 1(a)** depicts selected demographics for the selected study counties and non-Georgia.

**Table 1(a).**
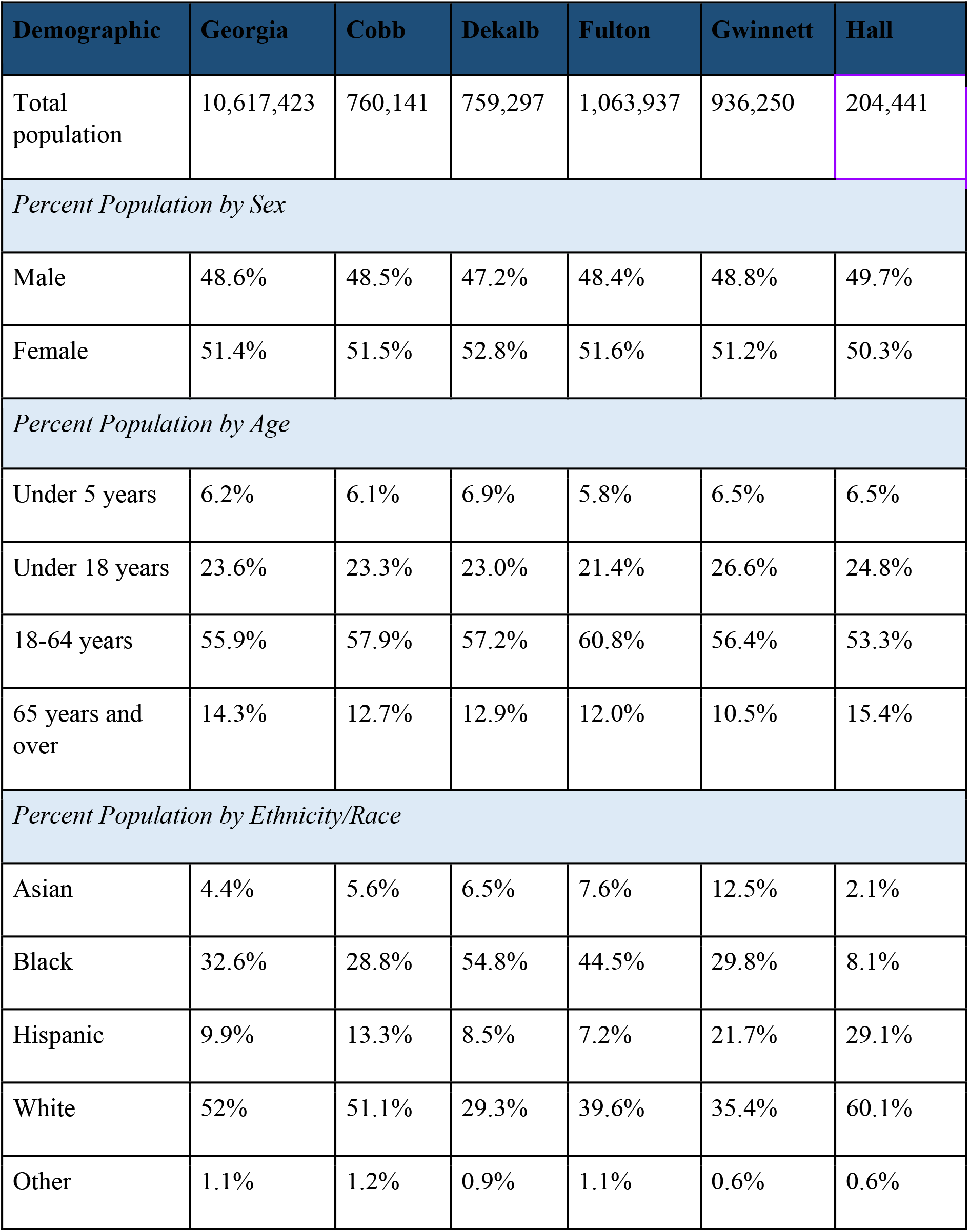
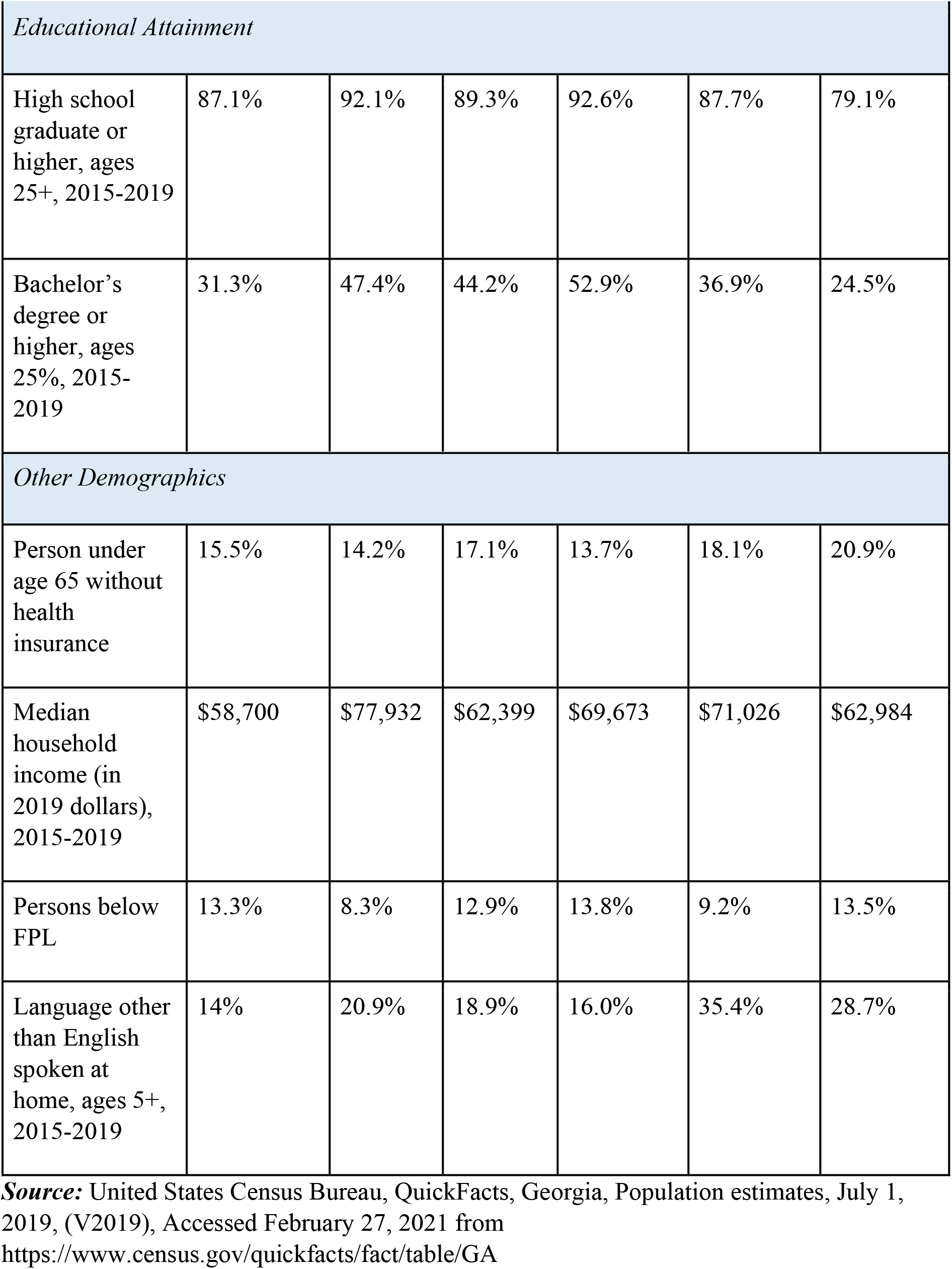
Selected demographics in Georgia and study counties.

| **Demographic** | **Georgia** | **Cobb** | **Dekalb** | **Fulton** | **Gwinnett** | **Hall** |
| --- | --- | --- | --- | --- | --- | --- |
| Total population | 10,617,423 | 760,141 | 759,297 | 1,063,937 | 936,250 | 204,441 |
| *Percent Population by Sex* |  |  |  |  |  |  |
| Male | 48.6% | 48.5% | 47.2% | 48.4% | 48.8% | 49.7% |
| Female | 51.4% | 51.5% | 52.8% | 51.6% | 51.2% | 50.3% |
| *Percent Population by Age* |  |  |  |  |  |  |
| Under 5 years | 6.2% | 6.1% | 6.9% | 5.8% | 6.5% | 6.5% |
| Under 18 years | 23.6% | 23.3% | 23.0% | 21.4% | 26.6% | 24.8% |
| 18-64 years | 55.9% | 57.9% | 57.2% | 60.8% | 56.4% | 53.3% |
| 65 years and over | 14.3% | 12.7% | 12.9% | 12.0% | 10.5% | 15.4% |
| *Percent Population by Ethnicity/Race* |  |  |  |  |  |  |
| Asian | 4.4% | 5.6% | 6.5% | 7.6% | 12.5% | 2.1% |
| Black | 32.6% | 28.8% | 54.8% | 44.5% | 29.8% | 8.1% |
| Hispanic | 9.9% | 13.3% | 8.5% | 7.2% | 21.7% | 29.1% |
| White | 52% | 51.1% | 29.3% | 39.6% | 35.4% | 60.1% |
| Other | 1.1% | 1.2% | 0.9% | 1.1% | 0.6% | 0.6% |

| *Educational Attainment* |  |  |  |  |  |  |
| --- | --- | --- | --- | --- | --- | --- |
| High school graduate or higher, ages 25+, 2015-2019 | 87.1% | 92.1% | 89.3% | 92.6% | 87.7% | 79.1% |
| Bachelor's degree or higher, ages 25+, 2015-2019 | 31.3% | 47.4% | 44.2% | 52.9% | 36.9% | 24.5% |
| *Other Demographics* |  |  |  |  |  |  |
| Person under age 65 without health insurance | 15.5% | 14.2% | 17.1% | 13.7% | 18.1% | 20.9% |
| Median household income (in 2019 dollars), 2015-2019 | \$58,700 | \$77,932 | \$62,399 | \$69,673 | \$71,026 | \$62,984 |
| Persons below FPL | 13.3% | 8.3% | 12.9% | 13.8% | 9.2% | 13.5% |
| Language other than English spoken at home, ages 5+, 2015-2019 | 14% | 20.9% | 18.9% | 16.0% | 35.4% | 28.7% |

**Source:** United States Census Bureau, QuickFacts, Georgia, Population estimates, July 1, 2019, (V2019), Accessed February 27, 2021 from https://www.census.gov/quickfacts/fact/table/GA

**Table 1(b).** Percentage of COVID-19 cases and deaths by selected ethnicity/race and sex.

| Measure | Cobb | Dekalb | Fulton | Gwinnett | Hall |
| --- | --- | --- | --- | --- | --- |
| *Ethnicity/Race\** |  |  |  |  |  |
| *American Indian/Alaska Native* |  |  |  |  |  |
| % of cases | % | % | % | % | % |
| % of deaths | 0.4% | 1.0% | - | - | - |
| % of population | 0.5% | 0.5% | 0.3% | 0.8% | 0.8% |
| *Asians* |  |  |  |  |  |
| % of cases | % | % | % | % | % |
| % of deaths | 2.4% | 5.6% | 1.6% | 8.8% | 1.1% |
| % of population | 5.6% | 6.5% | 7.6% | 12.5% | 2.1% |
| *Blacks* |  |  |  |  |  |
| % of cases | % | % | % | % | % |
| % of deaths | 43.2% | 60.9% | 67.8% | 30.0% | 8.2% |
| % of county population | 28.8% | 54.8% | 44.5% | 29.8% | 8.1% |
| *Native Hawaiian/Pacific Islander* |  |  |  |  |  |
| % of cases | % | % | % | % | % |
| % of deaths | 0.2% | 0.5% | - | 0.5% | % |
| % of county population | 0.1% | 0.1% | - | 0.1% | 0.2% |
| *Whites (includes Hispanics)* |  |  |  |  |  |
| % of cases | % | % | % | % | % |
| % of deaths | 45.3% | 27.8% | 28.8% | 51.0% | 86.4% |
| % of county population | 64.4% | 37.8% | 46.8% | 57.1% | 89.2% |
| *Other* |  |  |  |  |  |
| % of cases | % | % | % | % | % |
| % of deaths | 8.5% | 4.2% | 1.8% | 9.8% | 3.8% |
| % of county population | 1.2% | 0.9% | 1.1% | 0.6% | 0.6% |
| *Sex* |  |  |  |  |  |
| *Female* |  |  |  |  |  |
| % of cases | % | % | % | % | % |
| % of deaths | 51.7% | 43.6% | 48.7% | 58.7% | 48.4% |
| % of population | 51.5% | 52.8% | 51.6% | 51.2% | 50.3% |
| *Male* |  |  |  |  |  |
| % of cases | % | % | % | % | % |
| % of deaths | 48.3% | 56.4% | 51.3% | 41.3% | 51.6% |
| % of county population | 48.5% | 47.2% | 48.4% | 48.8% | 49.7% |

**Note:** \*excluded “unknown” race/ethnicity in total amounts for these calculation

As of February 26, 2021, each county had the following confirmed COVID-19 cases: 55,157 (Cobb); 51,483 (Dekalb); 74,050 (Fulton); 80,100 (Gwinnett); 23,758 (Hall); and 24,385 (non-GA).^7^ These counties top Georgia’s list of most confirmed cases and are mostly urban, with the exception of Hall County. However, these counties do not rank highest among the counts of cases per 100,000 population. In this regard, they are near Georgia’s average at around 6,500 cases per 100,000, with the exception of Hall County at 11,513.5 cases per 100,000 and Gwinnett County at 8,248 cases per 100,000.^7,8^

### Course of COVID-19 in Georgia

Fulton County documented the first two cases of COVID-19 in Georgia on March 2, 2020.^7,8^ Reported cases quickly followed in counties surrounding Fulton, specifically Cobb, Dekalb, Gwinnett, and Hall, in vastly higher numbers than counties outside the larger Atlanta metropolitan area.^7,8^ Georgia banned large group gatherings on March 24, 2020 and ordered shelter-in-place on April 03, 2020. By late April, every Georgian county had reported cases.^7,8^ On April 30, 2020, Georgia lifted its shelter-in-place order, and on June 01, 2020, certain businesses, schools, and events reopened as group-size restrictions loosened. While the number of total cases remained high in Fulton and surrounding counties, the cases per 100,000 were not high; these metropolitan counties stayed consistent with the Georgia average.^7,8^ Meanwhile, the cases per 100,000 people in rural counties surpassed the state average.^7,8^

Fulton, Cobb, Dekalb, Gwinnett, Hall, and non-GA counties reported the highest total confirmed deaths in Georgia as well. As of February 27, 2021, each county had the following number of deaths: 1,045 (Fulton), 838 (Cobb), 762 (Dekalb), 886 (Gwinnett), 373 (Hall), and 406 (non-GA).^7,8^ The disease continues to spread, with a total of 29,014 new cases in the last two weeks and an average rate of 2,072 new cases per day as of February 26, 2021^7,8^. On January 5, 2021, Georgia reported its first case of COVID-19 UK variant, new strain B.1.1.7, and the infected person was in isolation during contact tracing.^9^

### Sociodemographic differences among COVID-19 cases in Georgia

Studies from the earliest days of COVID-19 showed sociodemographic differences such as race, age, gender, and presence of chronic illness could affect the occurrence and disease severity. A study conducted March-April, 2020 with COVID-19 patients from six different Atlanta hospitals indicated the following factors were independently associated with hospitalization for COVID-19: age 65+, Black race, diabetes mellitus, obesity, smoking, and lack of insurance.^10^ Another study that occurred March-May, 2020 in rural southwest Georgia showed similar results; hypertension, age 65+, and obesity were independent predictors of mortality^11^. Of the 522 hospitalized participants in the study, 87% were Black, median age was 63 years, and 53% were females; males had a higher mortality rate than females by about 9%.

Despite the age group of 10-59 years constituting approximately 75% of confirmed cases in Georgia, they accounted for about 15% of deaths; patients ages 60+ comprise nearly 85% of confirmed deaths.^7,8^ A steadily increasing percentage of confirmed cases end up hospitalized with an increasing age group. There was approximately a 30% hospitalization rate for those aged 80 years and older, a 23% hospitalization rate for those aged 70-79 years, and a 13% hospitalization rate for those aged 60-69 years, with those 59 and younger trailing below 10%.^7,8^ When infected with SARS-CoV-2, some patients develop antibodies (seropositivity) while others do not. A study with infected participants from Dekalb and Fulton Counties in Georgia found adults aged 18-49 years and 50-64 years had the highest seroprevalence at 3.3% and 4.9%, respectively.^12^

Data from the urban Georgian counties of Fulton, Dekalb, Cobb, and Gwinnett; and the rural county of Dougherty found that children and adults under age 60 may be 2.38 times more infectious than adults over the age of 60.^13^ In another Georgia study, the transmission was most frequent between people of the same age group for all age groups.^14^ Transmission occurred more frequently between different age groups if a relationship existed (family, friend, coworker, etc.) as compared to transmission in public spaces.^14^ The same study found that the majority of transmission came from people aged 40-70 years at the very beginning of the pandemic but shifted to those aged 25-50 years in June-July, 2020.^14^

Males had a higher mortality rate than females in Georgia from COVID-19, with 23% mortality for males versus 13.8% for females.^11^ Males were also hospitalized for COVID-19 more than females, even after controlling for various other factors such as age and chronic conditions.^10^ However, more females than males have been infected overall, with females comprising 53.1% of confirmed cases and males 45.4%.^7,8^ Transmission from a primary male case-patient to a secondary male case-patient was much less frequent than their transmission to a secondary female case-patient. Still, primary female case-patients are equally transmitted to secondary males and females, which the study attributed to the role of females as caregivers.^14^ There was not much difference between males and females in building SARS-CoV-2 antibodies, with both measuring about 2.5% seroprevalence.^12^

Total cases in Georgia are highest among the Whites—about 260,000 cases, followed by Blacks-nearly 160,000, unknown ethnicity/race about 130,000, “other” races-over 43,000, and Asians-12,000.^7,8^ Among known cases, about 67,000 have been in Hispanics.^7,8^ The percentage of cases hospitalized was highest among Blacks at 10.9% followed by Whites at 7.6%.^7,8^ These results were consistent with studies that highlight the Black race as a risk factor for hospitalization.^10,11^ Counties with greater proportions of Black inhabitants reported more COVID-19 related deaths and cases even after adjusting for other socioeconomic factors.^16^ Blacks have shown a 5.2% seropositivity for antibodies compared to a 0.3% for non-Hispanic Whites.^12^ Studying the demographic breakdown of cases in a state as diverse as Georgia will help tailor educational materials and intervention approaches.

In this paper, the authors explored the demographic breakdown among COVID-19 cases in the six groups aforementioned. Comparisons were made by race/ethnicity, age, gender, and presence of chronic disease. With increased knowledge about which groups and areas COVID-19 impacts in Georgia, pandemic response teams can improve prevention measures, modify techniques of vaccine delivery, and refine health education for their target group.

This study examined whether gender-specific and racial/ethnic differences exist in GDPH COVID-19 data and obtained the predictive models and its summaries for both case and death occurrences. The objectives of this study were to: (i) detect the specific regions of Georgia that are most affected by COVID-19 and find any public health loopholes, advice, or policies that work for the future wellbeing of the residents; (ii) identify infected confirmed cases and deaths among males and females through descriptive analysis of accessible sociodemographic variables; (iii) carry out artificial neural network models to classify chronic disease conditions in deaths; (iv) develop a confusion matrix to visualize the performance of prediction; (v) depict the area under the curve with receiver operating characteristic (AUC-ROC) to measure the true positive rates against false positive rates; (vi) utilize Bayesian analysis to estimate posterior models and its inferences for cases and deaths; and (vii) describe the public health implications.

## Methodology

### Data source/variables and study population

The study utilized data reported for February 01 to November 10, 2020 from the GDPH website (https://dph.georgia.gov/covid-19-daily-status-report)(15). The GDPH released a limited number of de-identified COVID-19 data for both aggregate as well as individual levels. The study population included confirmed COVID-19 cases via PCR tests for six groups/counties: Cobb, Dekalb, Fulton, Gwinnett, Hall Counties, and non-Georgia residents identified by RGIS software^1^, **Figure 1(a)**. Study variables included age, gender, cases, deaths, counties, and chronic diseases/medical-conditions. The age variable was grouped into subgroups 0-17 years, 18-39 years, 40-59 years, 60-79 years, and ≥80-99 years. The gender was categorized by male and female; underlying medical conditions were classified by yes, no, unknown; and confirmed number of cases and deaths were extracted from 161 counties. Non-Georgian residents were considered as a group and consist of people visiting/commuting from one state to another state for their daily jobs or other activities.

**Figure 1(a).**
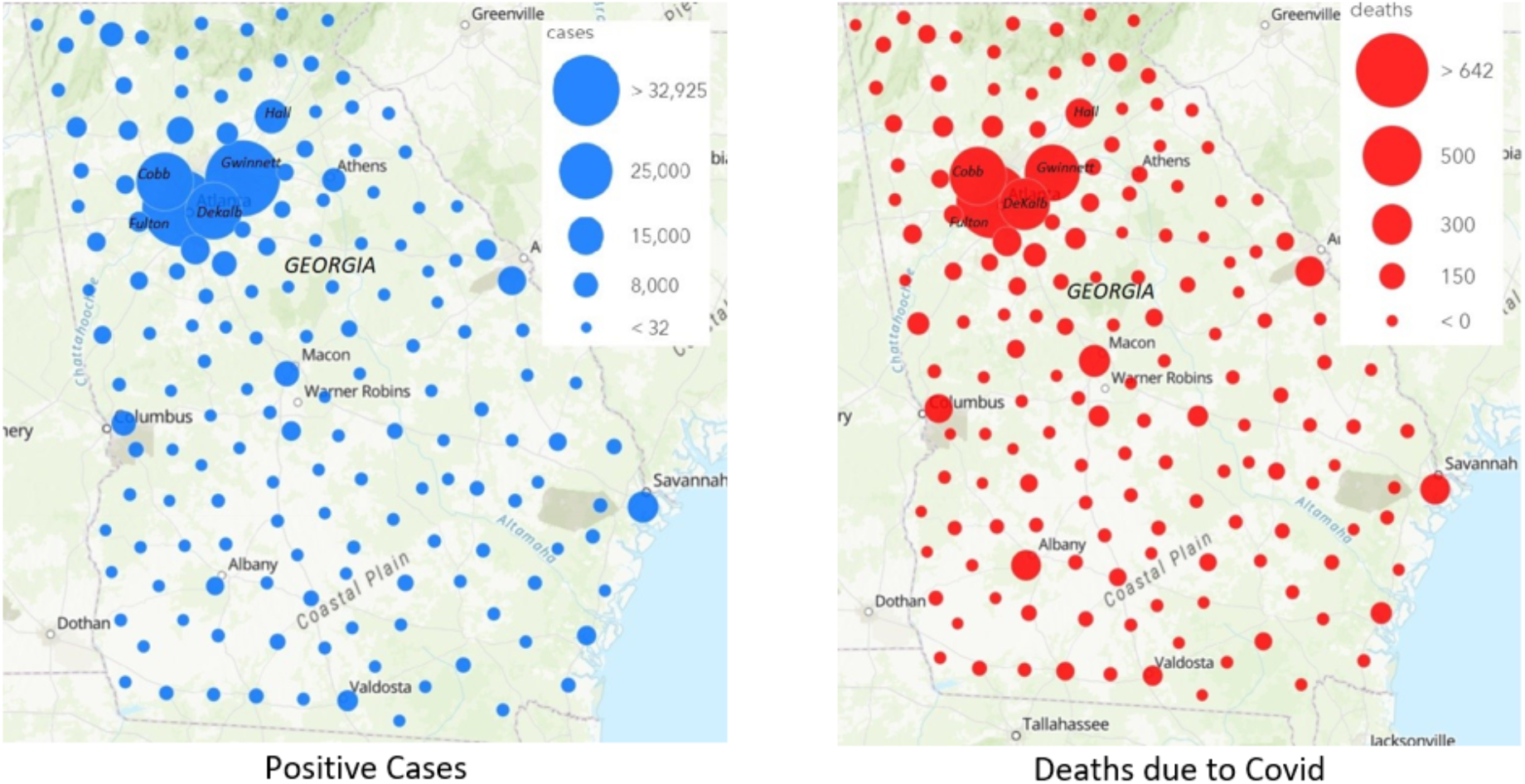
GIS map for cases and deaths and bubbles indicate intensities.

**Figure 1(b).**
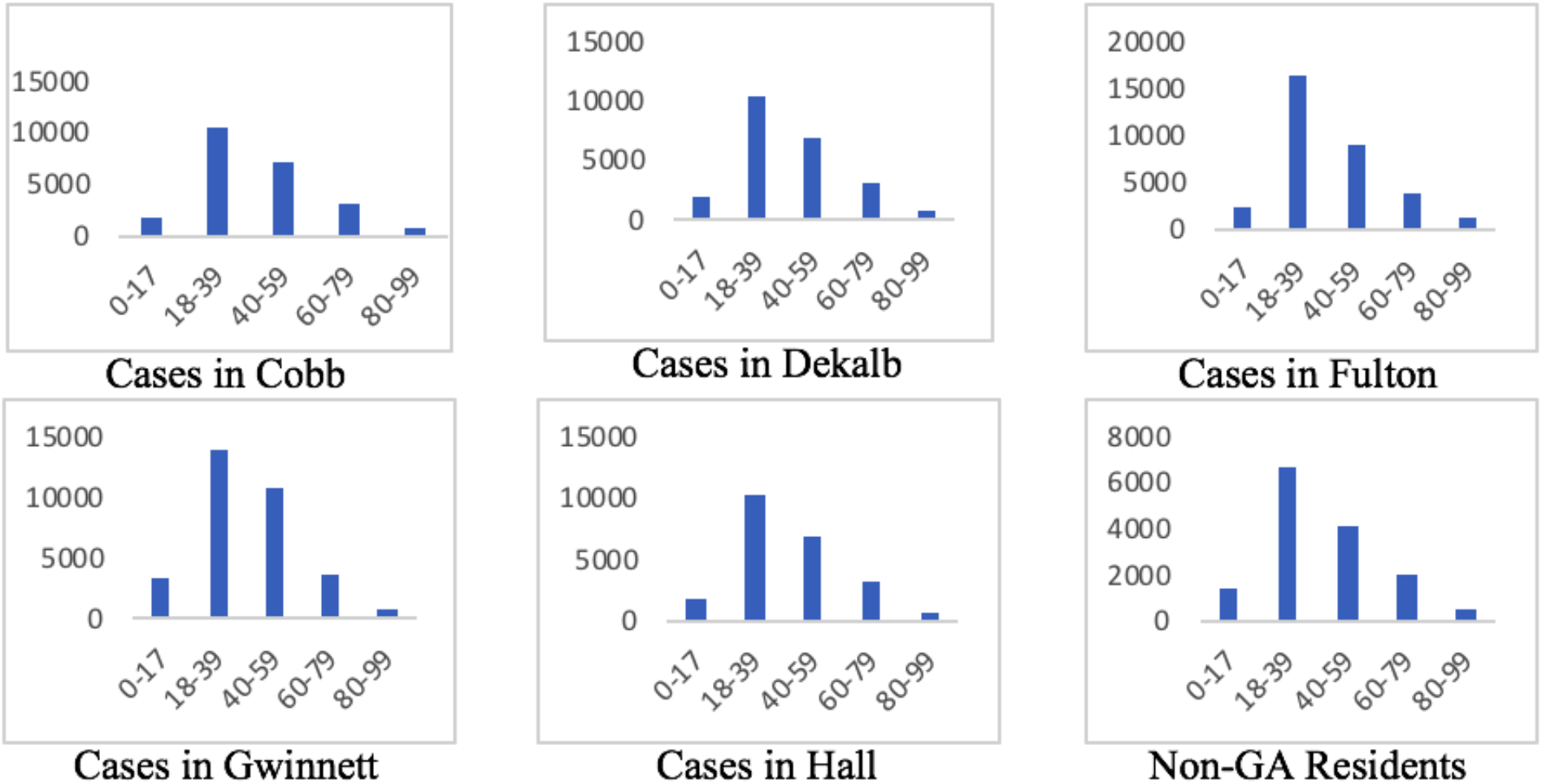
Bar diagram for COVID-19 positive cases for six counties.

**Figure 1(c).**
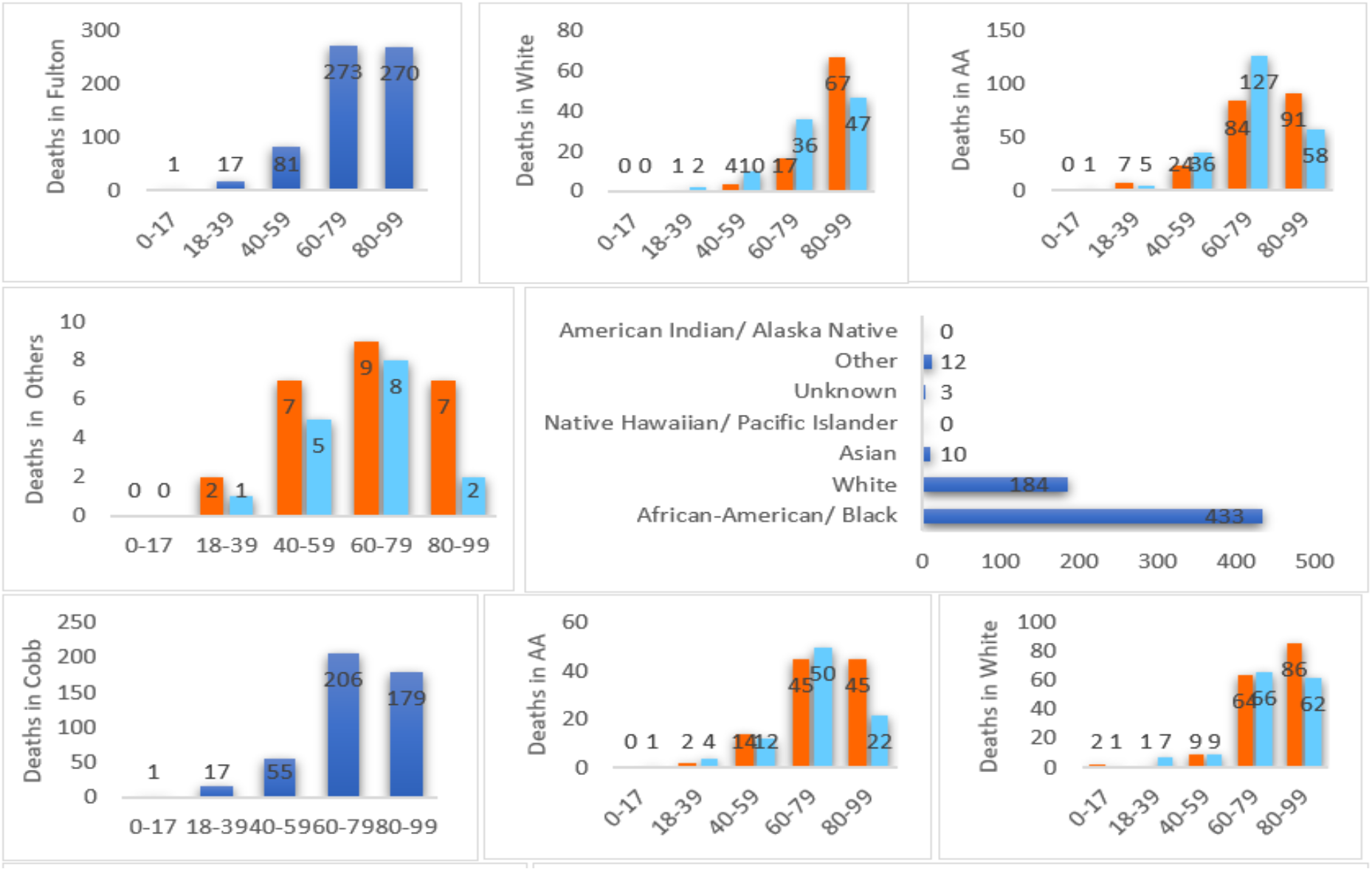
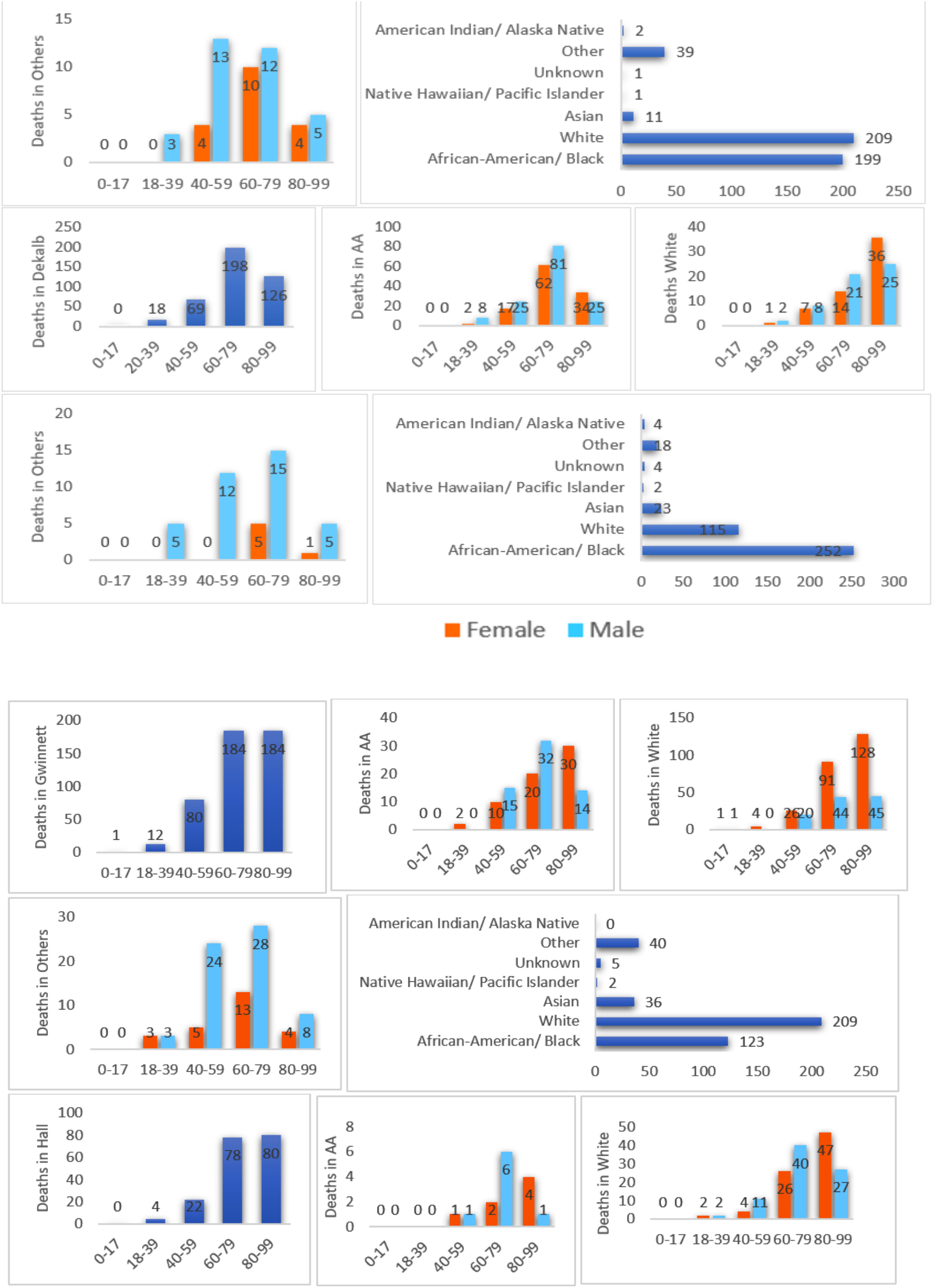
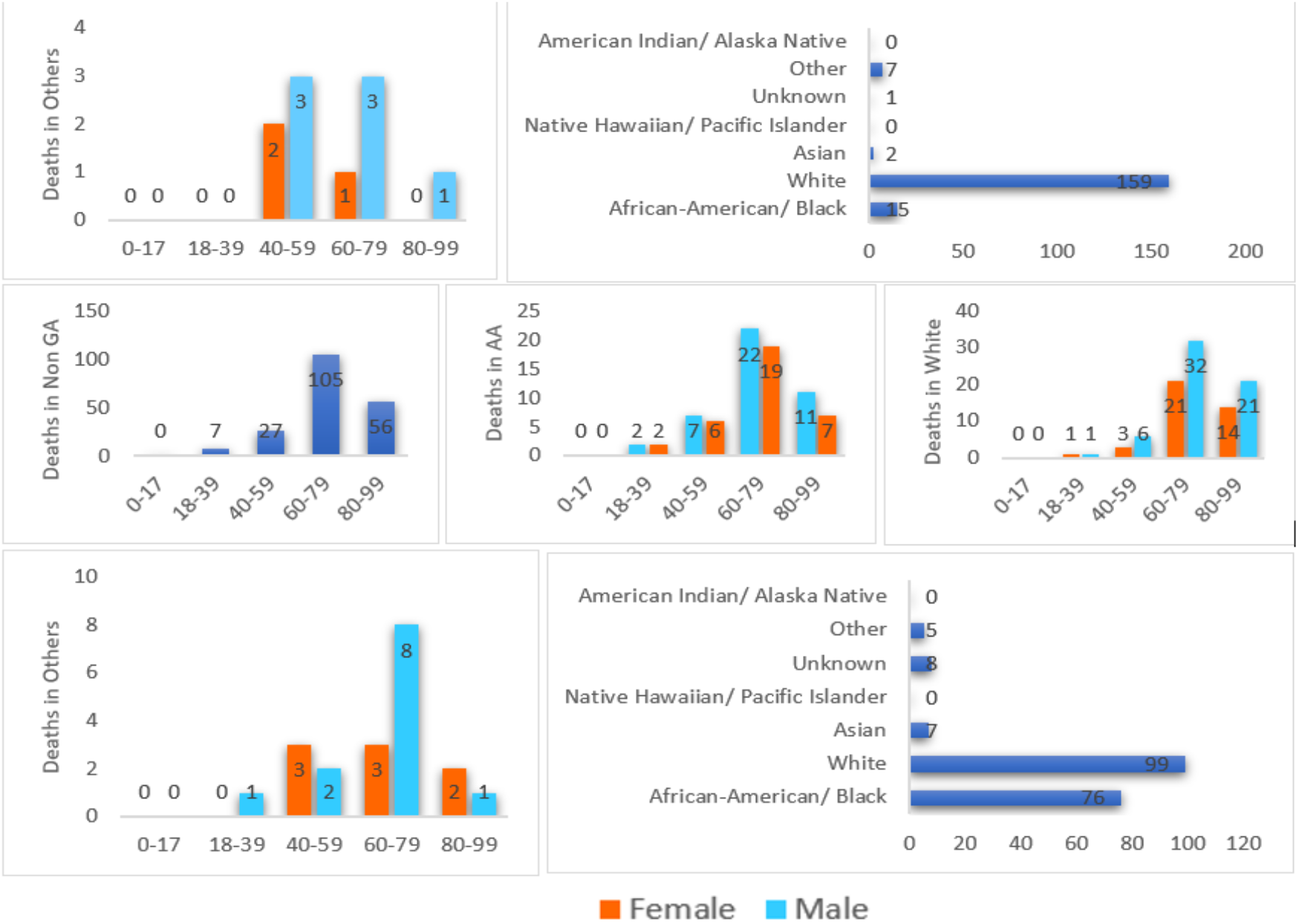
Bar diagram for deaths for six counties including non-GA residents.

### Exploratory data analysis

Two bubble maps (big/small) depicted the intensity of cases and deaths using the RGIS software. Each bubble indicates the intensity of positive cases and deaths due to COVID-19 based on the radius of the bubble. Note that big bubbles indicate more cases or deaths. Moreover, separate bar charts were displayed to show the comparisons of cases and deaths based on race and gender. The cases and deaths were distributed into five age groups according to the direction of the GDPH. The Kolmogorov-Smirnov and Shapiro-Wilk tests were used to check the data normality.

### Bayesian analysis

Bayesian inference is the process of drawing a logical conclusion about a population or probability distribution from data using Bayes’ theorem, which is the product of the prior for the parameter and the likelihood function for the data, to produce the posterior probability distribution, which is the conditional distribution of the uncertain future parameter given the data. For a scientific investigation, an experienced expert can derive a prior about the parameter that is an informed guess from personal experience, pilot study, or particular domain knowledge. The Bayesian Markov chain Monte Carlo^17^ is a useful method to obtain the posterior distributions or models of risk parameters by interfacing R on WinBUGS14 software. This method assumes the observed *data*, or ***z*** *=* {*z*_*1*_, *z*_*2*_, *…, z*_*n*_}, is normally distributed with mean (*μ*), unknown variance (*τ*), and its sampling distribution follows *z*∼*N*(*μ,τ*). The prior distributions for *μ* and *τ* are as follows: *μ*∼ *N*(*θ,τ, θ*). and *τ*∼*γ* (*α, β*); where 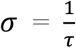. After a number of iterations using the WinBUGS14 and R software, one can simulate the estimates for the parameters *mu, tau, sigma*, and *theta*. A diffuse prior may be considered for *tau* and *theta* when the prior knowledge of them is unknown.

### Artificial neural network

For classification analysis using data science techniques (**Figure 2(d)**), the ANN method was used.^18,19,20^ Data set was split into training (70%) and testing (30%) sets by stratified random sampling (**Table 2(c)**). The ANN was implemented on training data. In the learning phase, the network learns by adjusting the weights to predict the correct class label of the given inputs. The following equation was used to classify chronic condition of deaths, 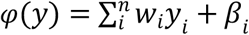 where *y*_1_, *y*_2_,…, *y*_*n*_ were the input variables, and *w*_1_, *w*_2_,…, *w*_*n*_were the respective weights. *β* was the bias, which was added to the weight inputs to form the net inputs accurately. Both bias and weights were adjustable parameters of the neurons of the ANN model.^21^ The ANN worked like neuron’s which acted like mathematical functions that classified the information. *φ*(*y*) was worked with sigmoid function 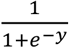 which helped to determine the class or labels. Here, *φ*^[1](*i*)^ was the output from *i*th neuron of the first layer. Therefore, *φ*^[1](*i*)^(*x*) = *w*^[1]^*y*^(*i*)^ + *β*^[1](*i*)^ passed through the tangent hyperbolic activation function to find *λ*^[1](*i*^)(*y*) = *tanh* (*φ*^[1](*i*)^(*y*)) Then, the output layer was *φ*^[2](*i*)^(*y*) = *w*^[2]^*λ*^[1](*i*)^(*y*) + *β*^[1](*i*)^. So, predictive probabilities were evaluated through 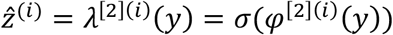. Based on probability, the predictive labels were determined. From this method, a confusion matrix was constructed to evaluate predictive classification of deaths with chronic conditions. Accuracy of the predictive model had been determined by the formula, *Accuracy =* (*TP+TN*)*/* (*TP+TN+FP+FN*); where TP, TN, FP, and FN were true positive, true negative, false positive and false negative, respectively. Using testing data sets, AUC-ROC^21,22^ was applied to visualize the performance of a classifier over all possible thresholds, obtained by graphing the false positive rate against the true positive rate as the threshold for assigning observation to a given class was varied.

**Table 2(a).** Summary statistics for posterior mean with 95% credible intervals based on deaths.

|  | Posterior Mean | Credible Interval |  | Posterior Mean | Credible Interval |  |
| --- | --- | --- | --- | --- | --- | --- |
|  | Female | 2.5% | 97.5% | Male | 2.5% | 97.5% |
| **Cobb** | 73.29 | 71.08 | 75.42 | 68.89 | 66.42 | 71.34 |
| **Dekalb** | 73.30 | 71.05 | 75.46 | 68.36 | 66.44 | 70.29 |
| **Fulton** | 74.34 | 72.37 | 76.40 | 73.48 | 72.18 | 74.49 |
| **Gwinnett** | 77.17 | 74.23 | 80.07 | 70.54 | 67.74 | 73.40 |
| **Hall** | 75.22 | 73.25 | 77.16 | 68.74 | 66.88 | 70.54 |
| **non-GA** | 70.28 | 67.22 | 73.31 | 70.96 | 68.48 | 73.31 |

**Table 2(b).** Summary statistics for posterior mean with 95% credible intervals for both genders.

|  | Posterior Mean | Credible Interval |  | Posterior Mean | Credible Interval |  |
| --- | --- | --- | --- | --- | --- | --- |
|  | Cases | 2.5% | 97.5% | Deaths | 2.5% | 97.5% |
| **Cobb** | 4641 | 4577.86 | 4701.91 | 93.41 | 41.91 | 144.61 |
| **Dekalb** | 4537 | 4474.17 | 4598.93 | 82.02 | 34.61 | 129.26 |
| **Fulton** | 6550 | 6488.14 | 6612.76 | 128.06 | 73.78 | 182.79 |
| **Gwinnett** | 6483 | 6421.54 | 6545.15 | 92.08 | 42.82 | 141.26 |
| **Hall** | 2022 | 1959.73 | 2083.72 | 36.89 | 3.49 | 70.92 |
| **non-GA** | 2963 | 2900.55 | 3024.81 | 38.98 | 3.17 | 74.13 |

**Table 2(c).** Case processing for all six counties using data science technique.

| Data splitting |  | Dekalb | Hall | Cobb | Gwinnett | non-GA | Fulton |
| --- | --- | --- | --- | --- | --- | --- | --- |
| Sample | Training | 281 | 120 | 316 | 314 | 143 | 457 |
|  | Testing | 130 | 57 | 142 | 147 | 52 | 145 |

**Table 2(d).** Confusion matrix for classifications.

|  | Dekalb |  |  |  | Cobb |  |  |  | Gwinnett |  |  |  |
| --- | --- | --- | --- | --- | --- | --- | --- | --- | --- | --- | --- | --- |
|  | 0 | 1 | 2 | PC | 0 | 1 | 2 | PC | 0 | 1 | 2 | PC |
| 0 | 1 | 29 | 0 | 3.3 | 0 | 22 | 0 | 0 | 0 | 24 | 0 | 0 |
| 1 | 0 | 91 | 0 | 100 | 0 | 99 | 0 | 100 | 0 | 112 | 0 | 100 |
| 2 | 0 | 9 | 0 | 0 | 0 | 6 | 0 | 0 | 0 | 11 | 0 | 0 |
| % | 0.8 | 99.2 | 0 | 70.8 | 0 | 100 | 0 | 78 | 0 | 100 | 0 | 76.2 |
|  | Hall |  |  |  | non-GA |  |  |  | Fulton |  |  |  |
|  | 0 | 1 | 2 | PC | 0 | 1 | 2 | PC | 0 | 1 | 2 | PC |
| 0 | 0 | 8 | 0 | 0 | 0 | 15 | 0 | 0 | 88 | 4 | 0 | 95.4 |
| 1 | 0 | 45 | 0 | 100 | 0 | 22 | 1 | 95.7 | 43 | 5 | 0 | 10.7 |
| 2 | 1 | 3 | 0 | 0 | 0 | 12 | 1 | 7.1 | 44 | 1 | 0 | 0 |
| % | 0 | 98.2 | 0 | 78.9 | 0 | 96.2 | 3.8 | 44.2 | 94.6 | 5.4 | 0 | 50.3 |

**Notes:**
- % sign means overall percentage correctly predicted. - PC means Percentage Correct. - Deep Orange colors indicate predicted classes and green colors indicate true classes.

**Figure 2(a).**
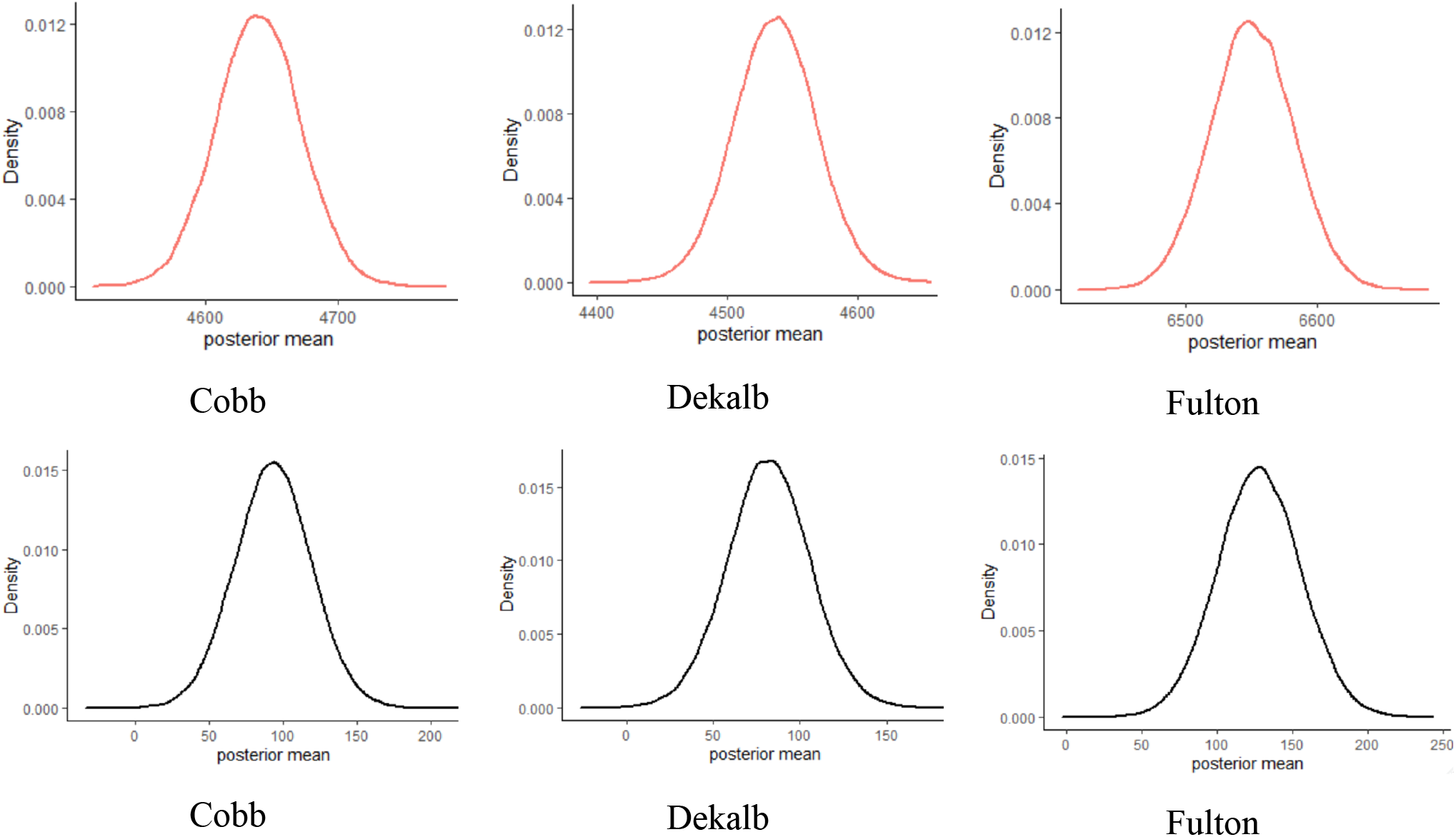
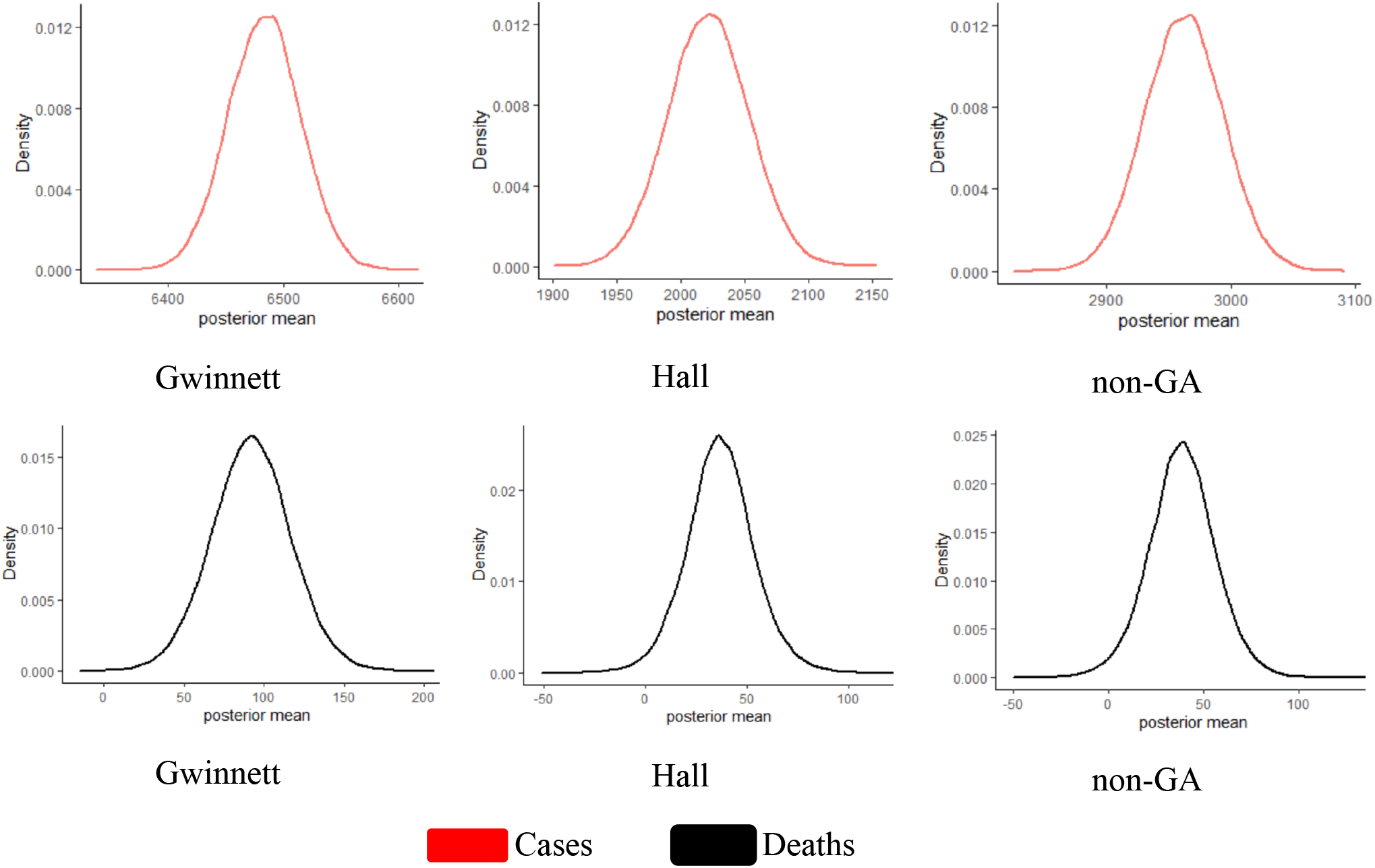
Posterior mean of cases and deaths.

**Figure 2(b).**
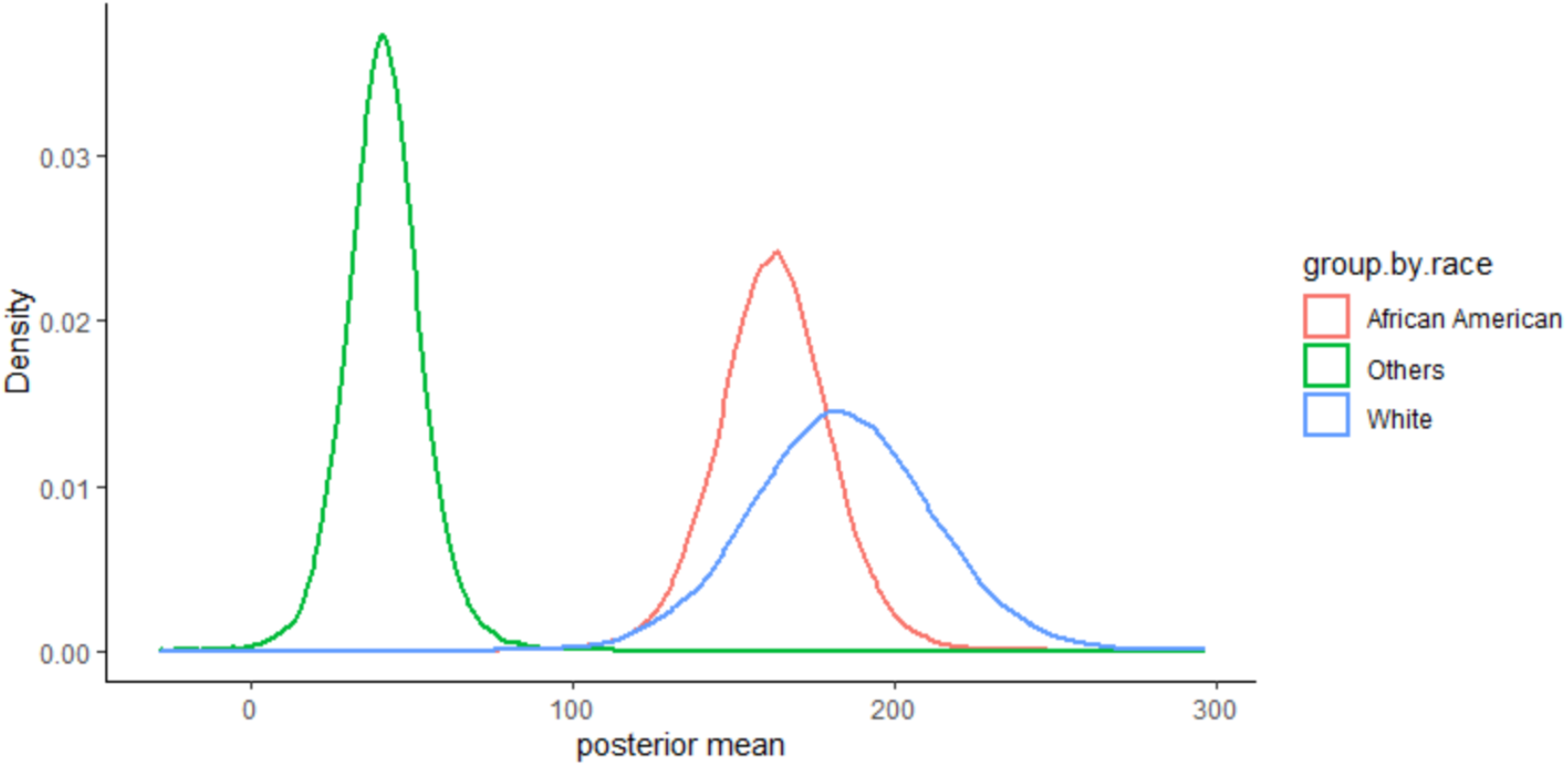
Posterior mean of deaths by race overall six counties.

**Figure 2(c).**
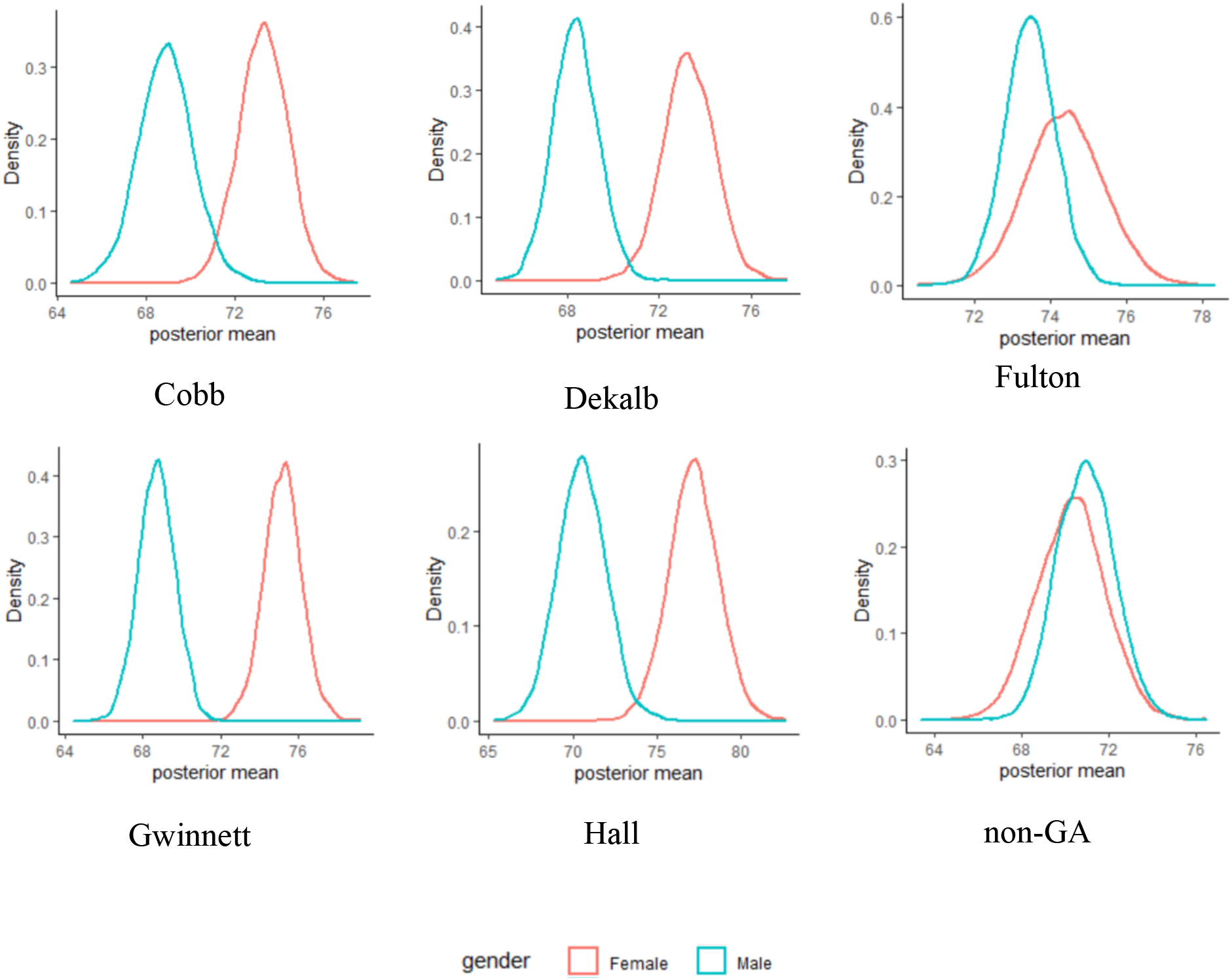
Posterior mean distribution for female and male in case of deaths.

**Figure 2(d).**
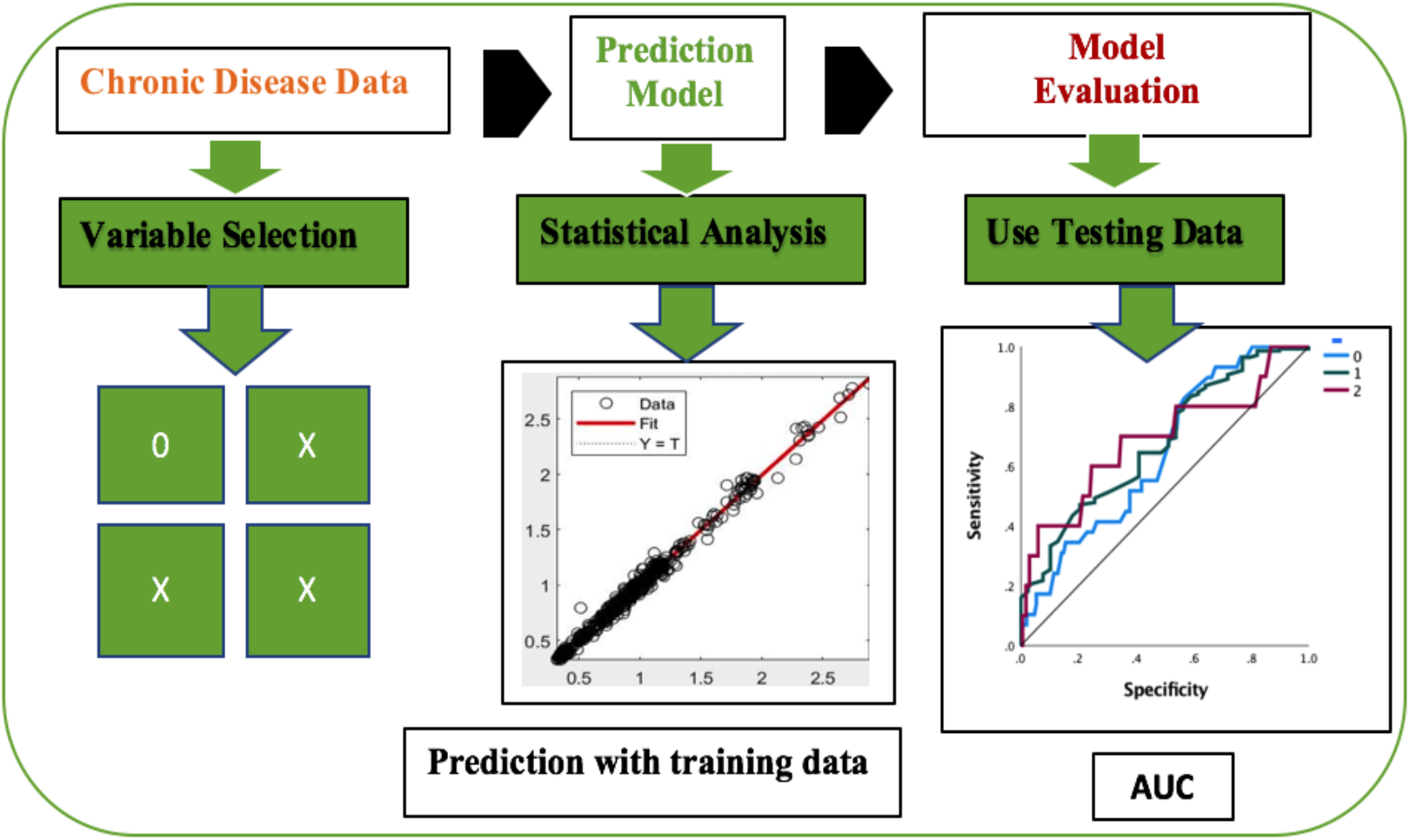
Architecture of statistical machine learning methodology.

**Figure 2(e).**
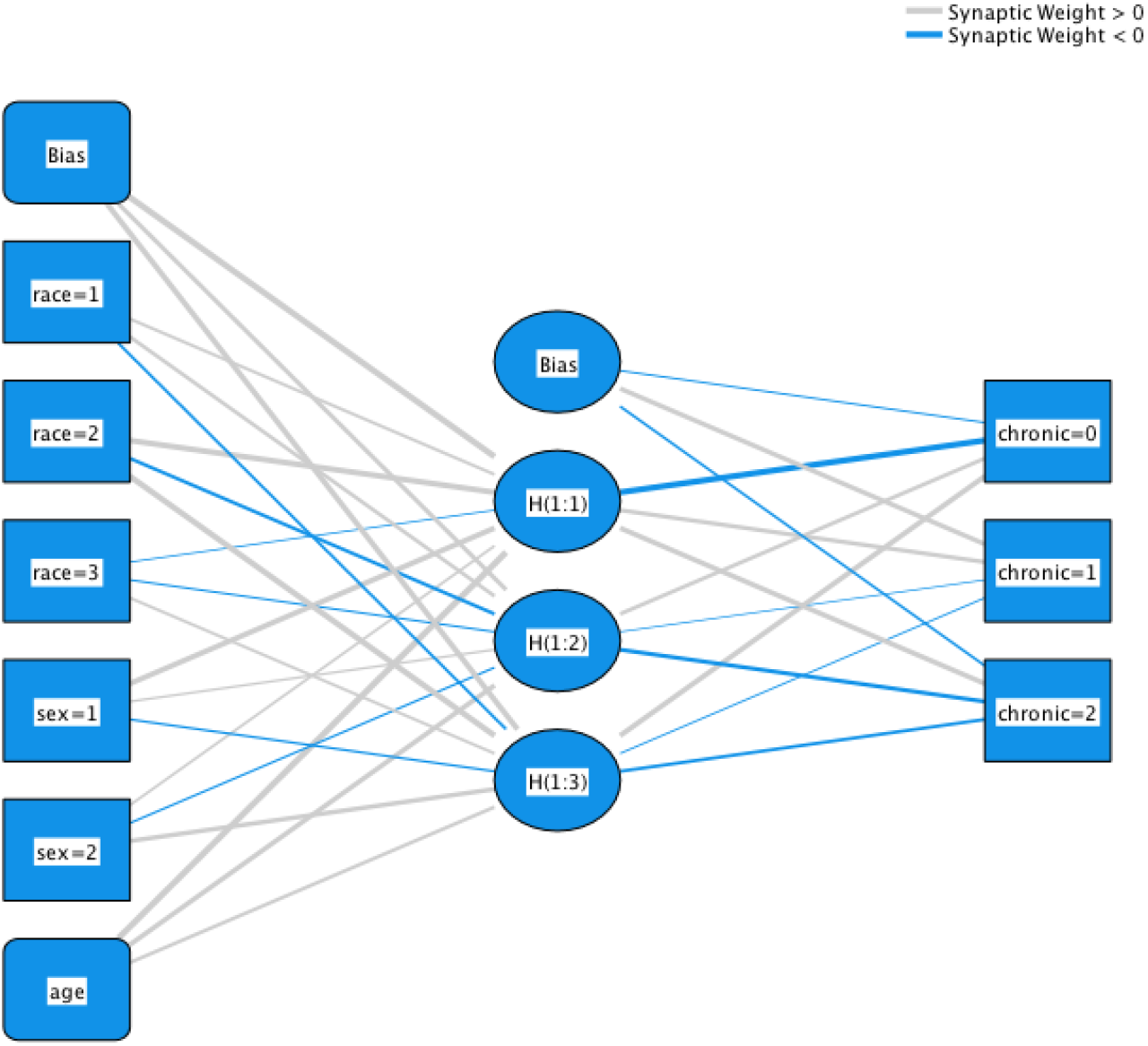
ANN architecture for classification of chronic conditions.

**Figure 2(f).**
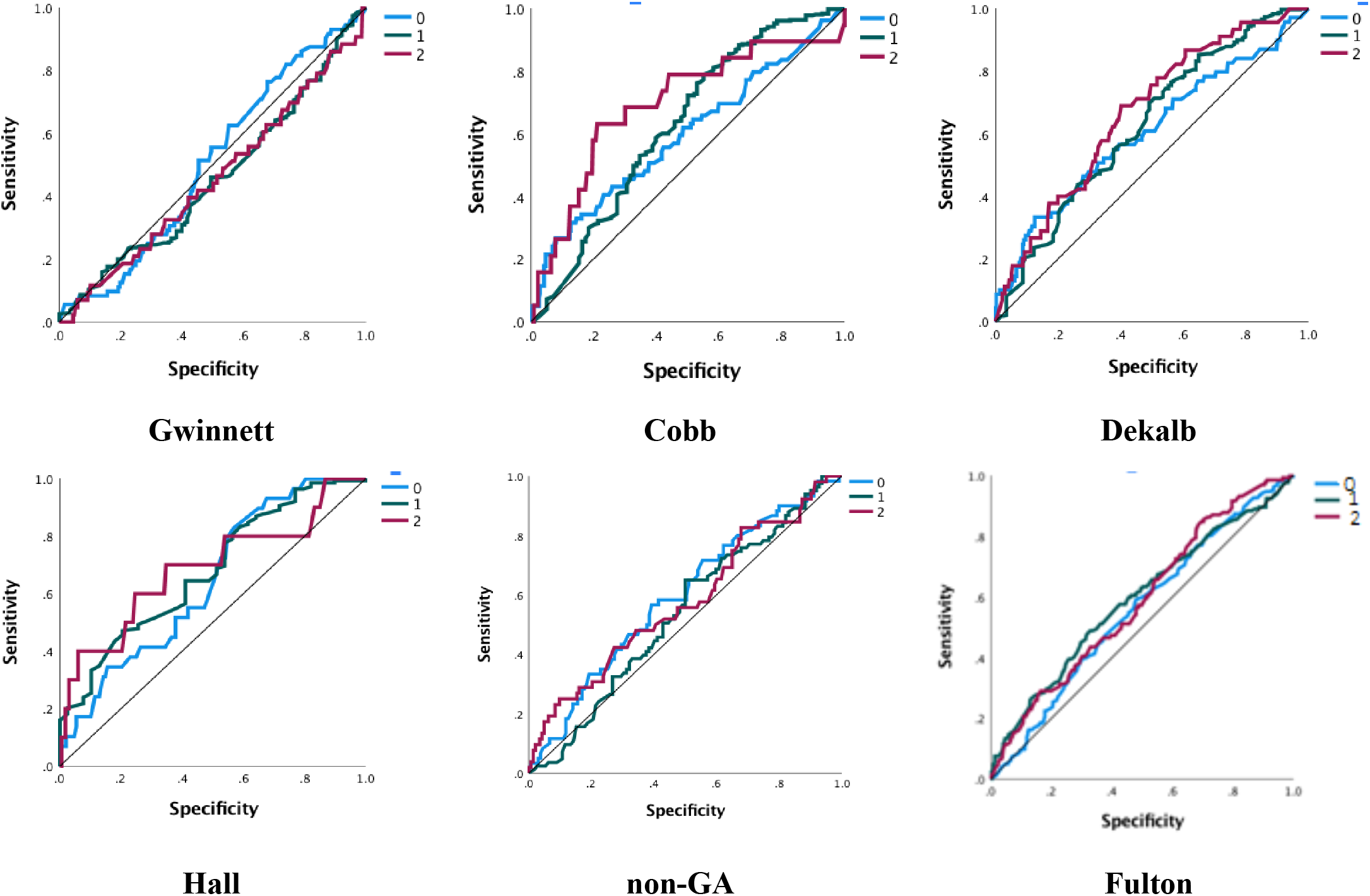
Sensitivity analysis of ANN models, 1-specificity versus sensitivity.

**Figure 2(g).**
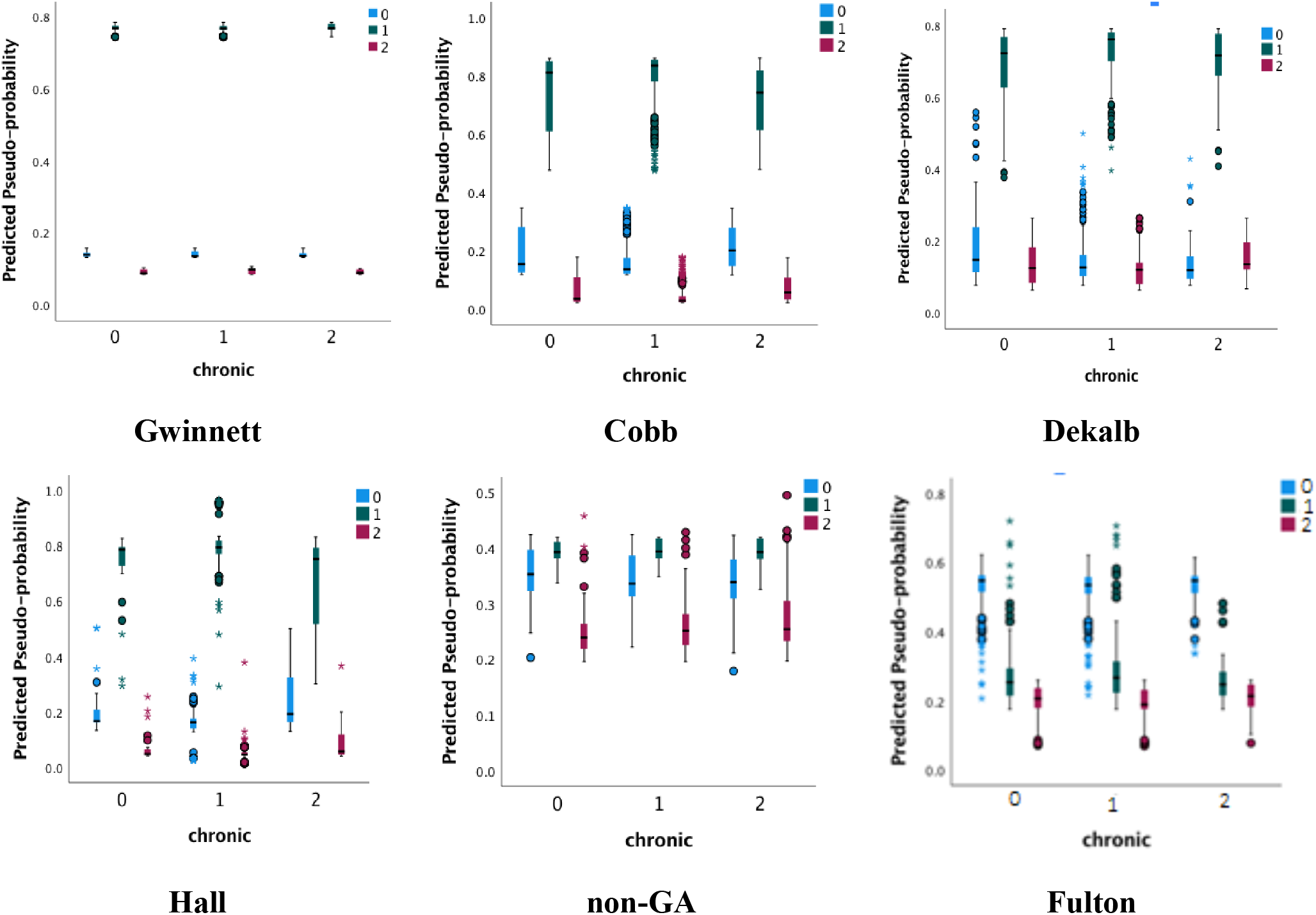
Chronic disease vs predicted pseudo probability plots for all six counties.

## Results and discussion

According to the bubble map by RGIS software, positive COVID-19 cases and consequent deaths were shown in some populated counties in the state of Georgia. The map displays five big bubbles (counties) amongst 161 bubbles (160 Georgia counties and a bubble for non-Georgia residents). The study included the non-Georgia resident group due to its significant impact. Selected bubbles were based on the highest number of COVID-19 positive cases and subsequent deaths. Selected bubbles also represented top populated counties (Fulton, Dekalb, Hall, Gwinnett, Cobb, respectively) in Georgia according to the U.S. Census Bureau. To show the deaths due to COVID-19, bar graphs were used to identify cases by age group and race. The variable “age” had five groups with a twenty-class width each for restricted ages under 99 years. Moreover, to present death counts, six ethnic/racial groups (African-Americans/Blacks, Whites, Asian, Native Hawaiian/Pacific Islander, Unknown, and Others) were shown by horizontal bar graphs.

Fulton County had the highest number of COVID-19 positive deaths and cases. Except for Hall County, all the others (Fulton, Cobb, Dekalb, and Gwinnett) were very close to the metropolitan city-Atlanta. **Figure 1(c)** exhibits, the overall number of deaths in older people was higher. The bar indicated an increasing number of deaths with increasing age. Both 60-79 and 80-99 age groups presented higher numbers of deaths (42.52% and 42.05%, respectively). A bar chart counts deaths and shows the variations based on ethnicity/race. Blacks had the highest number of deaths, almost 67.4% of total deaths due to COVID-19 positive cases. Whites accounted for 28.66% of the deaths, which was 38.74% less than Blacks. Asians comprised 10 percent deaths, which was relatively small compared to deaths in Blacks. Moreover, analysis investigated death counts based on sex in Whites, Blacks, and Other groups. The number of deaths in females was higher than males for Whites; however, Black males were more affected in Fulton County. The number of deaths increased with female age in other ethnic/racial groups. There was a significant difference in Black male cases; the number of deaths in Black males dropped 69% from the 60-79 to 80-99 age group. The rate of death in White females was higher than males, although the opposite situation occurred for Black males aged 60-79. In Cobb County, the highest number of deaths occurred in the 60-79 and 80-99 age groups. Most COVID-19 deaths occurred in older, White persons. There was only one recorded COVID-19 positive death in the 0-17 age group.

Considering deaths by ethnicity/race, Cobb, and Gwinnett Counties have similar results, opposite of those from Fulton and Dekalb Counties, where most deaths occurred in Blacks. Almost 45.23% of total deaths in Cobb County occurred in Whites. However, when examining death rate by sex and ethnicity/race, the scenario differs for Gwinnett County. More White males deaths occurred in 60-79 years compared to White females. Another intriguing situation of note was for Fulton County, almost 29.33% of deaths occurred in Blacks, which was the recorded highest death rate. Overall, patients in the 60-79 and 80-99 age groups had higher death rates compared to other age groups. However, most COVID-19 positive cases occurred in ages 18 to 39, which agrees with the study of Lau et al.^13^

The data for both cases and deaths were checked to see if they follow a specific probability distribution. By making use of the Kolmogorov-Smirnov and Shapiro-Wilk test, the data were verified and found to be normally distributed. It was noted that some data for the cases and deaths may follow normal distribution by the study of Dunn and Smyth.^23^ The Bayesian analysis predicts the future variability of deaths and cases in gender and race/ethnicity. These analyses were performed and the summary results for the posterior means in case of both genders and overall deaths for race/ethnicity were obtained. To derive the posterior distributions, we used Jeffreys prior^17^ since there were limited data for the variables. We reported the summary results for the Bayesian posterior model parameters given the database of the cases and deaths of COVID-19 patients by interfacing R-language on WinBUGS14 software. A data-based posterior distribution was generated after the sample was simulated 50,000 times, and posterior densities were displayed. These densities characterize the future trends of the prognosis of coronavirus development for the cases and deaths. The shape of the distribution of parameters (*mu* and *theta*) for gender in cases and deaths was normal, while the others (*tau* and *sigma*) were asymmetrical. **Table 2(a)** reflects the summary results of the posterior means in cases both genders. One can compare the point estimators and 95% credible intervals of the posterior mean for each county. **Table 2(a)** reveals the summary results for posterior means in overall cases and deaths. The distribution of location parameter *mu* was observed to be approximately normal and the distributions of precision parameters *sigma* and *tau* were exhibited as skewed to the right. The overall posterior distribution of deaths for race and ethnicity were displayed in **Figure 1(b)**, and it was obtained that Whites had the highest mean number of deaths compared to Blacks and Others. For overall race/ethnicity in five counties and non-GA, Blacks, Whites, and Others had the posterior means for deaths 162.48, 183.18, and 41.35, respectively, where the 95% credible intervals were 127.15–197.42, 128.29–238.27, and 17.97–66.14, respectively.

The ANN model was used to classify which deceased persons did or did not have chronic disease with the unknown class. The predictors were age, race, and sex, and the response was chronic disease conditions: yes, no and unknown. The ANN classification model generalized people who died from COVID-19 disease and also had chronic diseases. **Table 2(d)** describes actual versus predicted classification of three levels (0 = no, 1 = yes, 2 = unknown) for each county. The applied neural net model found high precision of class predictions. In Fulton County-which had the highest number of COVID-19 cases, the model delineated less performance with non-GA had the lowest between all, it also showed less accuracy. Overall accuracy of ANN output for the Cobb, Dekalb, Fulton, Gwinnett, Hall, and non-GA were 70%, 78%, 50%, 76%, 79%, and 45%, respectively. Results laid out a specificity versus sensitivity curve for the counties in **Figure 2(f)** with three chronic condition curves. The AUC-ROC depicted a good model diagnostic ability for each county except Gwinnett (below the diagonal line or chance level for groups 1 and 2). For each category, the predicted pseudo-probability in **Figure 2(g)** of being in that category was higher for a randomly chosen case in that category than for a randomly chosen case not in that category. For the categorical responses (no, yes, unknown), the predicted-by-observed chart displays clustered boxplots of predicted pseudo-probabilities for the combined training and testing data samples. The boxplot in **Figure 2(g)** with 0.5 and above probability on the vertical axis depicts the correct predictions, and below the probability 0.5 represents incorrect predictive classifications. Hall, Cobb, Gwinnett, Dekalb Counties represent good classification for predicting cases compared to non-GA and Fulton Counties.

### Public health implications

Posterior summary statistics indicate that more Whites died from COVID-19 compared to Blacks and Others, and Blacks were the second highest group for deaths in Fulton and Dekalb Counties. On average, older (aged ≥ 60) males were highly affected in most of the counties with Fulton, Dekalb and non-GA groups showing a spike. Some potential reasons for these health disparities documented in previous studies include lower income, lower educational attainment, cultural factors and language barriers, lack of health insurance, lack of access to healthcare services, and lower quality healthcare services.^24,25,26,27^ As depicted in **Table 1(a)**, the counties seem to have health-related disparities. For example, rates for residents under age 65 without health insurance ranged from 13.7% (Fulton County) to 20.9% (Hall County). Persons below the Federal Poverty Level (FPL) ranged from 8.3% (Cobb County) to 13.8% (Fulton County). Language other than English spoken at home ranged from 16% (Fulton County) to 35.4% (Gwinnett County). Bachelor degree or higher ranged from 47.4% (Cobb County) to 24.5% (Hall County). Given the findings from this study that predict populations at greater risk and documented disparities, these counties may need to revise the annual health policies to improve the public health facilities.

The study aimed to increase knowledge of factors influencing COVID-19 transmission and disease severity in Georgia in order to develop gender-specific future virus models for effective interventions—including prevention measures; vaccine education, outreach and delivery; health education; and public health policy planning and implementation.

Predictive modeling on COVID-19 contributes to improvement of the existing public health policies, which are needed in light of the fact that differences in growth rates of COVID-19 have largely resulted from disparities in local public health policies (mask mandates, social distancing, etc.), individual and social behaviors, patient response, and more.^28,29^ The characteristics of future models help public health practitioners and researchers to understand the course of the COVID-19 pandemic and in turn provide guidance to help public health officials know how to prepare and anticipate transmission patterns and disease severity among diverse populations.^29,30,31^ For example, for the five counties in this study, public health officials can use information to decide where and whom to focus prevention, education, and vaccine efforts on to reduce transmission and minimize disease severity. However, predictive models will also create the need for more mathematical epidemiologists to manage outbreak dynamics and the need for more rapid, up-to-date data from public health and healthcare systems to better inform the application of effective, timely control measures.^32,33^

In closing, using the modelling strategies to predict the course of COVID-19 has critical implications for guiding public health policy making. The findings of this study will expand to identify infected individuals or counties for interventions for future pandemics. As data emerges from the pandemic, research teams will be better able to develop more robust models, with the end result of reducing spread and improving health outcomes.

### Conclusions

COVID-19 killed many more men than women where some suggested biological and social factors are associated.^34^ There were not enough significantly studied causes available in the published literature, but one possibility is that males received less education than females and may not get routine health screenings like women do. This result is consistent with the study of Shah and Killerby.^10,11^ Although it varies from state to state, some findings showed Whites died more compared to Blacks, proportionally due to more Whites living in those counties than other ethnic groups.^35^ At older ages, patients usually develop slower and weaker immune systems. Vaccines and flu shots may be less effective in developing an active, lasting response. Exercise and good nutrition may boost the immune system of infected COVID-19 patients. According to this study, most of the COVID-19 deaths were due to comorbidities related to chronic diseases. Many chronic diseases predispose individuals to have a less robust response against COVID-19, leading to severe outcomes such as death when faced with the additional burden of a respiratory illness. However, the type of chronic disease was unknown. Gwinnett, Fulton, and Cobb were the three counties with the highest death rates because these were the most populous counties. In Gwinnett County, there was a lack of nursing home staff such as nurses, doctors and healthcare workers.^36^ On the other hand, all of the counties were very close to Atlanta Metropolitan city, so the total and incoming population^37^ was very high. The ratio of physicians^38^ to patients was not enough for most of these counties. There was not sufficient data available for the count of nurses, but there were insufficient numbers of nursing homes. All of the counties had high living costs and median household income was around $69,000-$77,000. In addition to the high prevalence of comorbidities, some patients under 65 years of age did not have any health insurance, which can be assumed from the discussed **Table 1(a)**. Dekalb, Hall, and non-GA had less deaths compared to other counties, most likely because they are less populated than other three counties. In Dekalb, more Blacks died, whereas more Whites died in Hall county. The foremost reason was a smaller population of Black persons in Hall county. Whites died more than any other racial group in most counties studied, except Dekalb and Fulton counties, because most counties had more Whites in population; **Table 1(b)**. According to the results of our study, patients died more at ages over 59. There were some spikes found in most of the counties. No particular evidence or data had been found, but patients over 59 had more medical history such as chronic disease. Using Bayesian analysis, the distribution of posterior mean deaths for overall clearly indicated that the mean number of Whites died higher than Blacks, and Other groups. Although the percentage of the population of males was lower than females, death rates were higher in males. In most of the counties, male posterior mean deaths were higher than females, except for non-GA, which may indicate that a higher proportion of males suffered from chronic diseases compared to females. The ANN data driven scientific tools help to screen better predictive classification of chronic diseases in deaths and the AUC-ROC supports its validation.

A systematic review on modeling techniques for COVID-19 concluded that a larger dataset occurring over longer periods of time provided more accurate predictions than a smaller dataset for the cumulative deaths; Bayesian models appeared to provide more accurate and precise predictions.^29^ The use of large public health data, especially biological specimens, will be extremely valuable in developing a biomarker for outcomes research, quality assurance, public health surveillance, and other beneficial purposes. As noted in other predictive modeling studies on COVID-19, there are some challenges and limitations of predictive models. Data obtained early on in epidemics may be limited due to under-detection of cases, inconsistent detection of cases, reporting delays, and poor documentation–all of which affect the quality of model output.^39^ This is particularly true of the COVID-19 pandemic, where numerous cases were asymptomatic, leading to varying hypotheses as to the true prevalence of COVID-19 due to undetected cases.^40^ Predictive models lead to the possible exclusion of undocumented, unconfirmed cases due to lack of access to medical care, which could then lead to under-representation of some of the most vulnerable populations.^41,42^ Another limitation may be potential bias due to factors such as the accuracy of diagnostic tests, lasting immunity, reinfections, and population characteristics.^39^ The limitations of the present study are now stated as follows: he comorbidities of the chronic diseases were unknown because the data were only reflected with yes, no, and unknown; the GDPH did not release any gender specific data for cases and chronic diseases for race/ethnicity; and there were a very limited number of unidentified datasets publicly available. The study findings will help for interventions and develop policy briefs for future of pandemic, and they can be generalized to the population with geographic and gender-and race-specific similarities.

## Data Availability

The study utilized data reported for February 01 to November 10, 2020 from the GDPH website (https://dph.georgia.gov/covid-19-daily-status-report)(15). The GDPH released a limited number of de-identified COVID-19 data for both aggregates as well as individual levels.

## Acknowledgements

The authors would like to thank the Georgia Department of Public Health for releasing limited data to the public. This study was not financially supported.

## Footnotes

1 RGIS software, https://www.esri.com/en-us/arcgis/products/mapping

